# DOES QUALITY OF COUNSELING AND EXPOSURE TO FAMILY PLANNING MESSAGES INFLUENCE THE USE OF MODERN CONTRACEPTIVES AMONG WOMEN IN NORTHERN NIGERIA?

**DOI:** 10.1101/2024.04.03.24305214

**Authors:** Matthew Alabi, Leanne Dougherty, Eno-Obong Etim, Adebola Adedimeji

## Abstract

**Background:** Family Planning Counselling (FPC) involves information exchange on contraceptive methods between a provider and client and providing appropriate support in choosing a method that best suits the client’s needs. Access to sexual and reproductive health information enables women to make informed health decisions. However, the low modern contraceptive prevalence among women in northern Nigeria may be attributed to poor FPC and exposure to FP messages, among other factors. This study examines the impact of quality FPC and exposure to FP messages on modern contraceptive use.

**Methods:** We conducted a cross-sectional survey of 676 family planning clients aged 15-49 from 163 health facilities in Northern Nigeria. Quantitative data were collected using client exit interviews. The analysis included a binary logistic regression to predict the odds of modern contraceptive use using MII Plus and other explanatory variables.

**Result:** Only 29% received quality counselling about methods, while 35% were exposed to FP messages. The quality of counselling assessed using MII Plus did not predict modern contraceptive use. However, higher odds of using modern contraceptives were found among clients who reported their provider asked about their previous family planning experience (aOR=3.81; 95% CI: 1.04-13.99) and explained how the selected FP method works (aOR=5.51; 95% CI: 1.11-27.30). Clients who received FP messages from a place of worship (aOR=11.97; 95% CI: 1.76-81.62) and a community ruler (aOR=6.52; 95% CI: 1.02-41.58) had higher odds of using modern contraceptives.

**Conclusion:** Improving the quality of FPC and expanding exposure to FP messages through effective community structure can enhance the uptake of modern contraceptives in northern Nigeria.

## Background

Trends in modern contraceptive use over the past decade in Nigeria suggest a slow pace, with a Modern Contraceptive Prevalence Rate (mCPR) of 10.5% in 2011, 10.8% in 2016, and 18.3% in 2021(1). The 2022 estimate of 18% is considerably lower than the Federal Government of Nigeria’s target of 27% mCPR by the year 2020, despite various efforts to improve uptake including training of health workers and personnel on postpartum family planning, and implementation of task-shifting/sharing policy for essential healthcare services, among others(2). Besides the low national mCPR, there are regional differentials, with the highest mCPR of 21% in the southwest region, followed by the southeast (15%), north-central (12%), northeast 4% and northwest (3%) all lower than the national target(2).

Studies^3-11^ that have explored factors associated with modern contraceptive use among women, acknowledged the significant influence of socioeconomic variables and health-seeking behaviour. Studies(3–8) have also highlighted the influence of contextual factors and the mixed effects of fertility behaviour fueled by cultural norms and religious beliefs, autonomy, misconceptions, and fear of side effects as important predictors of modern contraceptives. However, there is evidence that Quality of Counseling (QoC) is associated with contraceptive utilization, patient outcomes, and reproductive health-seeking behaviour(9–13). The importance of quality counseling has received renewed interest necessitating the development of appropriate measures for assessing quality counseling. Examples of approaches for measuring QoC include process quality, which measures the exchange of information and interpersonal relationships that occur between the provider and client during care(14). Another measure is the Balanced Counseling Strategy (BCS), developed by the Population Council in the late 1990s to adapt counseling to an individual client’s specific reproductive health needs and enable the individual to select a method that best satisfies their specific needs(15,16). All these measures have been positively associated with contraceptive intention and continuation among users(17,18).

Recently, researchers have focused on using the Method Information Index (MII), and by extension, the MII Plus to assess the quality of counselling received by family planning clients. These measures are positively associated with contraceptive continuation regardless of the method(17,19,20). The MII is computed among current contraceptive users based on their responses to three questions, which ask whether the client received information on other contraceptive methods other than the one currently in use, whether clients were told about any possible side effects from using the current method, and whether they were provided with information on what to do should they experience any side effects (21). The MII Plus included one additional question that asks whether the client was told about the possibility of switching to another method should the method selected is deemed unsuitable (14). Each question requires a “yes” or “no” response. Those responding yes to all three or four questions are considered to have received adequate counseling, otherwise, counselling is considered inadequate(22).

Besides, studies(23–29) have also explored the association between exposure to family planning (FP) messages and use of modern contraceptives. Evidence from these studies suggest exposure to FP messages via different conventional and social media channels, was associated with a higher likelihood of modern contraceptive use. Although, few studies (24,30) have reported negative effects. Also, studies(4,9–11,13,23,31) have highlighted how religious and traditional systems could shape the influence of the mass media.

The MII has been adopted as one of the core measures for family planning 2020 initiative and constitute the primary indicator that addresses the issue of counselling, informed choice, and process quality (20). To our knowledge, no study has examined the influence of quality family planning counselling, and exposure to family planning messages on modern contraceptives use, and whether or not they predict usage in the northern regions of Nigeria where evidence suggests low uptake of modern contraceptives. The objective of this study is to bridge the gap in literature by examining the quality of client-provider interaction and exposure to family planning messages on modern contraceptive use..

## MATERIAL AND METHODS

### Study Setting

The study was conducted in six northern Nigerian states: Kano, Kaduna, Sokoto (Northwest), and Bauchi, Borno, and Yobe (Northeast). These states were purposively selected due to their participation in the Routine Immunization (RI) and, by extension Primary Health Care (PHC) Memorandum of Understanding (MoU) implementation (https://pdf.usaid.gov/pdf_docs/PA00WCCT.pdf). The RI/PHC MOU was borne out of the desire to address the insufficient and unpredictable funding for RI and PHC services, and the suboptimal commitment from the government to healthcare, by addressing issues relating to ownership and management in northern Nigeria associated with low RI coverage and poor PHC indicators.

### Study Design and Sampling

We conducted a cross-sectional health facility assessment with client exit interview and used a two-stage stratified sampling procedure in selecting the facilities. In the first stage, all LGAs in the RI implementation states have three (3) senatorial districts each. In ensuring coverage and spread in the evaluation states, the list of all the LGAs in each senatorial district was generated. On the assumption that implementation covers all the LGAs, fifty percent of the total number of LGAs in each state was selected proportionately (with exception of Borno). Although the evaluation LGAs were randomly selected, field realities such as security and access largely informed the final LGAs selected and surveyed as in the case of Borno. In each of the randomly selected LGAs, two PHCs were randomly selected and accessed to evaluate the efficiency and overall impact of the MoU.

The targeted number of health facilities across all the LGAs in all the MoU states was 156, at two health facilities per LGA. In selecting health facilities, consideration was given to location (rural/urban) except for cases where health facilities were considered not accessible for security reasons and distance. A total of 163 Health Facility Assessments (HFA) were conducted across all the six MoU states. The additional seven health facilities were because of oversampling in Borno state. Twenty family planning clients were targeted per health facility in all the states. Hence, a total of 676 Client Exit Interviews (CEI) were conducted with family planning clients across all six states. Clients were selected to participate in the exit interview if they met the inclusion criteria, namely married women in the age group 15-49 years and accessing family planning services at the health facility.

### Data Collection

Data collection was conducted concurrently across all six states between 14^th^ March and 8^th^ April 2022 following a pilot to assess the quality of the questionnaire. The questionnaire was programmed on mobile tablets using the SurveyCTO platform. The questionnaire, which was was translated into Hausa, the commonly spoken language in the study area, was adapted from the [WHO Service Availability and Readiness Assessment (SARA) Questionnaire](https://cdn.who.int/media/docs/default-source/service-availability-and-readinessassessment(sara)/sara_reference_manual_chapter2.pdf?sfvrsn=e46072be_3&ua=1). Eligibility criteria included women, ages 15-49, willing to provide informed consent after being provided with information about the study.

### Measurement of Variables

The primary outcome variable for this study is the use of modern contraceptives. The operational definitions for this and other selected explanatory variables are presented in Table 1. The selection of variables was largely informed by the review of the literature.

**Table 1:** Showing the operational definition of outcome and the explanatory variables used.

| Variables | Operational Definitions |
| --- | --- |
| *Outcome Variable* |  |
| Use of modern contraceptives | The outcome variable is the use of any modern contraceptives among the family planning clients coded as yes=1 and no=0.<br>Methods were further categorized as: Pills, barriers (male and female condoms), injectables, and device methods (implant and intrauterine device) |
| *Individual level variables* |  |
| Age of respondents | Self-reported age of the respondents as at survey time, categorized as: 15-24, 25-34, and 35-49 |
| Level of Education | Highest level of education attained: none, primary, secondary, and tertiary |
| Occupation | Current occupation: housewife/not working, self-employed, and formal employment |
| Distance to the health facility | Self-reported, categorized as: <10 minutes, 10-19, and at least 20 minutes |
| Autonomy | Primary decision maker on the use of modern contraceptives: Self, partner alone, or joint decision |
| Quality of Counselling (provider-client interaction) | Method Information Index Plus (MII Plus), an updated version of the MII. It consists of four (4) questions namely:<br>i. Whether the woman received information from the provider on the different family methods available.<br>ii. Provider talked about possible side effects associated with methods.<br>iii. Provider talked about what to do if the client experiences any side effects with the method selected.<br>iv. Provider talked about the possibility of switching to another method if the selected method is not suitable.<br>Each of the questions has a dichotomous response, yes coded as 1 and no, coded as 0. |
| Other counselling and communication measures | Provider used a visual aid to explain method; asked question about client previous FP experience; talked to the client about how the selected method work; told client when to return for follow-up; asked the client to fill a form with details of consultation; and asked if the client has any difficulty negotiating contraceptive use with her partner. Each is coded as yes=1, no=0. |
| Exposure to FP messages | Ever heard or seen messages on family planning in the last month preceding the survey (yes=1, no=0) |
| Sources of information on family planning | Heard or saw messages on family planning from community leaders, places of worship, and a health provider. Each option is coded as yes=1 and no=0. |

### Statistical Analysis

Both descriptive and inferential analyses were performed. The descriptive analysis performed includes frequency count, percentages, means, and standard deviations. For inferential analysis, logistic regression was used to check for relationships among variables. The dichotomous nature of the outcome variable influenced the conduct of binary logistic regression. Variables included in the adjusted multivariate model were chosen based on their contribution to the model and literature review. We presented the results of the analysis as odds ratios (ORs) and 95% confidence intervals. The goodness of fit of the model was used to determine if the application of multivariate logistic regression followed the required assumptions and whether the model sufficiently fit the data. The Hosmer-Lemeshow test, which tests for goodness of fit for the logistic regression model, was used, with a small chi-square and large p-value indicating a good fit to the data (χ^2^=354.79, p>0.05(p=0.653). The analysis of the data was performed using Stata version 14 software.

### Ethical Approval and Informed Consent

The study sought and received approval from the Population Council Institutional Review Board (Protocol Number 992). In Nigeria, ethical approval to conduct the study was obtained at the national and state levels. At the national level, approval was granted by the National Health Research Ethics Committee with approval number NHREC/01/01/2007-17/01/2022, while the State Health and Research Ethics Board in each of the study sites provided approval at the state level. Informed consent was presented to the study participants to read and either sign or thumbprint to give their consent. Thus, only caregivers who gave their consent were interviewed.

## RESULTS

### Description of Study Participants

Table 2 presents the profiles of the study participants. Participants were aged 16-49 years (mean 30), one quarter had no formal education, and nearly two-thirds were working, and more than half are literate with about 70% completing primary education or higher. One in four women reported reproductive decisions, including the use of contraception, are made by partners only.

**Table 2:** Socio-demographic Characteristics.

| **Maternal Characteristics** | **[N=676]** | **%** |
| --- | --- | --- |
| **State of Residence** |  |  |
| Bauchi | 120 | 17.7 |
| Borno | 121 | 18.0 |
| Kaduna | 140 | 20.7 |
| Kano | 144 | 21.3 |
| Sokoto | 84 | 12.4 |
| Yobe | 67 | 9.9 |
| **Region of residence** |  |  |
| Northeast | 308 | 45.6 |
| Northwest | 368 | 54.4 |
| **Caregivers age** |  |  |
| 15-24 years | 131 | 19.4 |
| 25-34 years | 344 | 50.9 |
| 35-49 years | 201 | 29.7 |
| **Level of Education** |  |  |
| No formal education | 206 | 30.5 |
| Primary | 99 | 16.6 |
| Secondary | 227 | 33.6 |
| Tertiary | 144 | 21.3 |
| **Literacy** |  |  |
| Cannot read and write | 247 | 36.6 |
| Can read and write | 376 | 55.6 |
| Can read or write | 53 | 7.8 |
| **Occupation** |  |  |
| Housewife/not working | 253 | 37.4 |
| Working | 423 | 62.6 |
| **Distance to health facility** |  |  |
| Less than 10 minutes | 164 | 24.3 |
| 10-19 minutes | 210 | 31.0 |
| At least 20 minutes | 302 | 44.7 |
| **Primary decision maker on contraceptive use** |  |  |
| Partner | 171 | 25.3 |
| Respondent | 68 | 10.0 |
| Joint decision | 437 | 64.6 |

### Quality of Counselling and Exposure to family planning Messages

As shown in Table 3, less than one-third (29%) of the women received adequate counseling, suggesting the quality of counselling was generally low. Assessment of other counselling indicators also revealed that only 28% reported the provider made use of visual aids during the counseling, while 32% reported their provider asked questions relating to family planning history and their preference for contraceptive methods. About 17% have heard or were exposed to family planning messages in the last 30 days preceding the survey.

**Table 3:** Client-Provider Interaction and Exposure to Family planning messages.

| **CLIENT-PROVIDER INTERACTION** | **[N=676]** |  |
| --- | --- | --- |
| **Method Information Index Plus** | **f** | **%** |
| Yes | 199 | 29.4 |
| No | 477 | 70.6 |
| **MII plus Questions** |  |  |
| Received information about the family planning methods |  |  |
| Yes | 228 | 33.7 |
| No | 448 | 66.3 |
| Received information about possible side effects associated with methods |  |  |
| Yes | 211 | 31.2 |
| No | 465 | 68.8 |
| The client received information on what to do if they experience any side effects with methods selected |  |  |
| Yes | 219 | 32.4 |
| No | 457 | 67.6 |
| The client received information on the possibility of switching to another method if the selected method is not suitable |  |  |
| Yes | 224 | 33.1 |
| No | 452 | 66.9 |
| **Other counselling and communication variables** |  |  |
| Provider used visual aid to explain how a method work |  |  |
| Yes | 186 | 27.5 |
| No | 490 | 72.5 |
| The provider asked about previous family planning experience |  |  |
| Yes | 215 | 31.8 |
| No | 461 | 68.2 |
| Provider told client when to return for follow-up |  |  |
| Yes | 232 | 34.3 |
| No | 444 | 65.7 |
| The provider asked if the client has challenge negotiating contraceptive use with a partner |  |  |
| Yes | 201 | 29.7 |
| No | 475 | 70.3 |
| The provider talked about how selected method work |  |  |
| Yes | 178 | 26.3 |
| No | 498 | 73.7 |
| **EXPOSURE TO FAMILY PLANNING MESSAGES** |  |  |
| Exposed to family planning information on media |  |  |
| Yes | 112 | 16.6 |
| No | 564 | 83.4 |
| Community leader as a source of information on family planning |  |  |
| Yes | 25 | 3.7 |
| No | 651 | 96.3 |
| Place of worship as a source of information on family planning |  |  |
| Yes | 24 | 3.6 |
| No | 652 | 96.4 |
| Health workers as a source of information on family planning |  |  |
| Yes | 72 | 10.7 |
| No | 604 | 89.3 |

**Table 4:** Relationship between sociodemographic factors, quality of FPC, media exposure to family planning messages and use of modern contraceptives.

| Variables | Outcome: use of modern contraceptives |  |  |  |
| --- | --- | --- | --- | --- |
|  | Unadjusted OR | 95% C.I. | Adjusted OR | 95% C.I. |
| **Region** |  |  |  |  |
| Northeast | 1.000 |  | 1.000 |  |
| Northwest | 0.52** | 0.36-0.76 | 0.33* | 0.15-0.73 |
| **Age Group** |  |  |  |  |
| 15-24 years | 1.000 |  | 1.000 |  |
| 25-34 years | 1.75* | 1.11-2.75 | 1.40 | 0.64-3.09 |
| 35-49 years | 1.67* | 1.02-2.76 | 1.45 | 0.61-3.48 |
| **Level of Education** |  |  |  |  |
| No formal education | 1.000 |  | 1.000 |  |
| Primary | 0.80 | 1.04-1.66 | 0.43 | 0.14-1.30 |
| Secondary | 0.70 | 1.35-1.98 | 0.62 | 0.17-2.32 |
| Tertiary | 1.06 | 2.26-3.66 | 1.30 | 0.29-5.69 |
| **Literacy** |  |  |  |  |
| Cannot read and write | 1.000 |  | 1.000 |  |
| Can read and write | 0.86 | 0.59-1.26 | 1.31 | 0.41-4.21 |
| Can read or write | 2.24 | 0.91-5.52 | 4.35* | 1.03-18.36 |
| **Work status** |  |  |  |  |
| Not currently working | 1.000 |  | 1.000 |  |
| Working | 1.69** | 1.14-2.50 | 2.17* | 1.06-4.46 |
| **Distance to a health facility** |  |  |  |  |
| Less than 10 minutes | 1.000 |  | 1.000 |  |
| 10-19 minutes | 0.52* | 0.31-0.87 | 0.75 | 0.32-1.75 |
| > 20 minutes | 0.59* | 0.36-0.97 | 1.46 | 0.64-3.35 |
| **Primary decision maker on contraceptives** |  |  |  |  |
| Partner | 1.000 |  | 1.000 |  |
| Wife | 2.60* | 1.15-5.86 | 1.88 | 0.59-5.93 |
| Joint | 1.12 | 0.75-1.69 | 2.95** | 1.41-6.17 |
| **Method Information Index Plus** |  |  |  |  |
| No | 1.000 |  | 1.000 |  |
| Yes | 0.24** | 0.16-0.35 | 0.40 | 0.13-1.23 |
| **The provider used a visual aid to explain method** |  |  |  |  |
| No | 1.000 |  | 1.000 |  |
| Yes | 0.32** | 0.22-0.47 | 2.44 | 0.92-6.48 |
| **The provider asked about previous FP experience** |  |  |  |  |
| No | 1.000 |  | 1.000 |  |
| Yes | 0.28** | 0.20-0.41 | 3.82* | 1.04-13.99 |
| **Provider told client when to return for follow-up** |  |  |  |  |
| No | 1.000 |  | 1.000 |  |
| Yes | 0.20** | 0.14-0.29 | 0.06* | 0.01-0.34 |
| **The provider asked if the client has a challenge negotiating contraceptive use with her partner** |  |  |  |  |
| No | 1.000 |  | 1.000 |  |
| Yes | 0.29** | 0.20-0.430 | 0.70 | 0.23-2.14 |
| **The provider talked about how selected method work** |  |  |  |  |
| No | 1.000 |  | 1.000 |  |
| Yes | 0.26** | 0.18-0.37 | 5.51* | 1.11-27.30 |
| **Media exposure to family planning messages** |  |  |  |  |
| No | 1.000 |  | 1.000 |  |
| Yes | 0.203** | 0.13-0.31 | 1.08 | 0.42-2.79 |
| **Place of worship as a source of information on FP** |  |  |  |  |
| No | 1.000 |  | 1.000 |  |
| Yes | 1.51** | 0.51-4.48 | 11.97* | 1.76-81.62 |
| **Community leader as a source of information on FP** |  |  |  |  |
| No | 1.000 |  | 1.000 |  |
| Yes | 2.23** | 0.66-7.57 | 6.52* | 1.02-41.58 |
| **Health worker as a source of information on FP** |  |  |  |  |
| No | 1.000 |  | 1.000 |  |
| Yes | 0.19** | 0.11-0.31 | 0.26* | 0.08-0.78 |

**\*\*significant as $p < 0.01$ ; \*significant at $p < 0.05$ , OR= odds ratio, aOR=adjusted odds ratio**

### Prevalence and Pattern of contraceptive use

The result of contraceptives prevalence is presented in Figure 1. Among the participants, injectables (42%), was the most commonly used method, followed by IUD and implant, accounting for (18%).

**Fig. 1:**
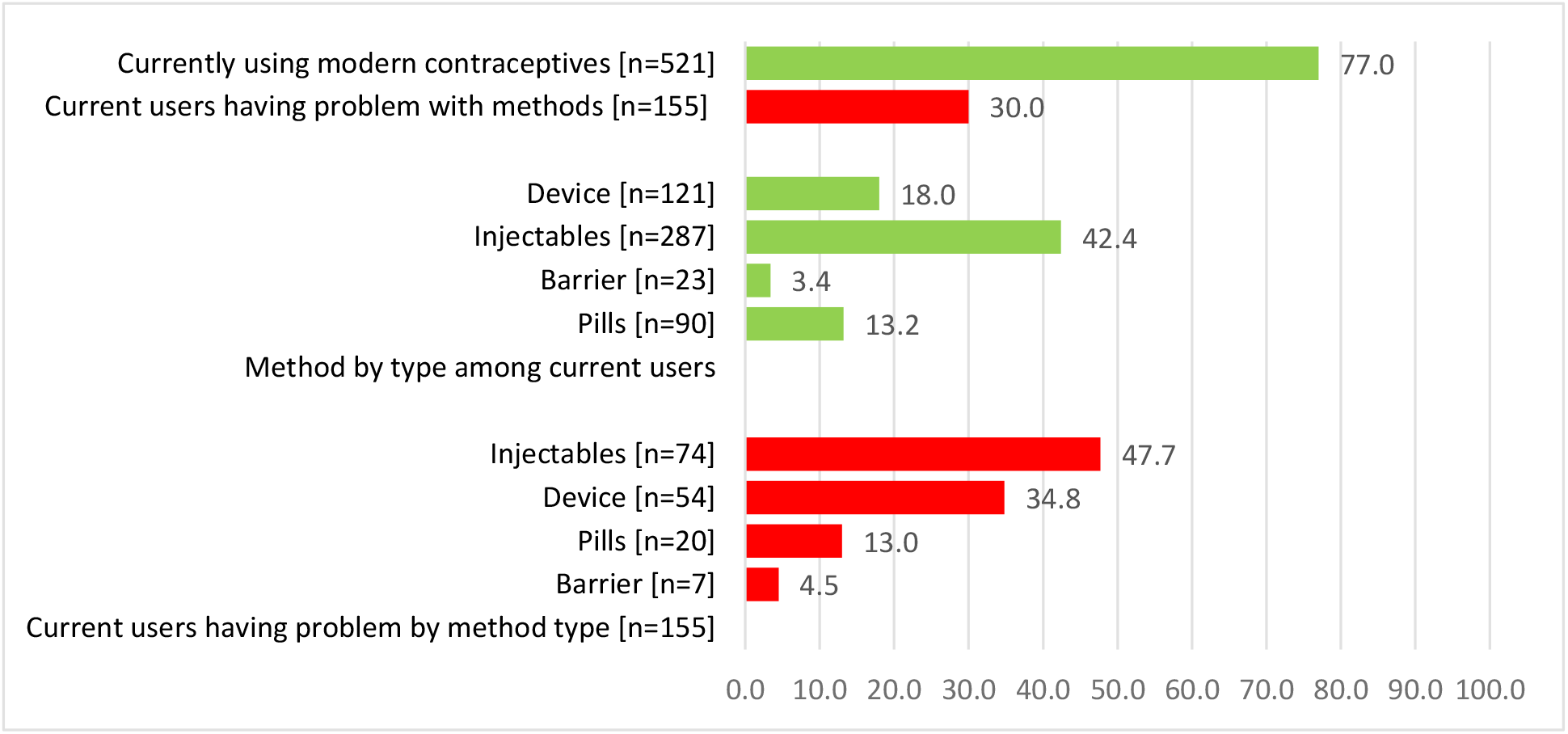
Pattern of Contraceptive Use among family planning clients.

### Multivariate Analysis

We performed a binary logistic regression analysis to determine factors influencing contraceptive use. At the individual level, region of residence, literacy level, being in employment, and decision-making autonomy were significantly associated with the use of modern contraceptives. Women residing in the northwest region (aOR=0.33 95% CI: 0.15-0.73) were less likely to use contraceptives relative to their counterpart in the northeast. Literate participants had higher odds of using contraceptives compared to those who are not literate (aOR=4.35 95% CI: 1.03-18.36). Those in employment were twice more likely to use contraceptives compared to unemployed women (aOR=2.17; 95% CI: 1.06-4.46). Caregivers with the ability to jointly make decisions regarding contraceptive use with their partner were nearly 3 times more likely to use contraceptives than those who have no autonomy to make decisions

### Quality of Family Planning Counselling (FPC)

MII Plus was the principal explanatory variable used to assess quality of FPC in this study, besides other measures. However, while the effect of MII Plus was not statistically significant in predicting contraceptive use (p>0.05), the effect of some other client-provider interaction and communication variables was statistically significant. For instance, clients who reported that their FP provider asked questions about their previous family planning experience had higher odds (aOR=3.82 95% CI: 1.04-13.99) of using contraceptives relative to women who reported no. Also, clients who reported their providers talked about how the selected method works (aOR=5.51; 95% CI: 1.11-27.30) were more likely to use contraceptives. However, clients who reported their provider told them when to return for follow-up showed lower odds (aOR=0.06; 95% CI: 0.01-0.34) of using contraceptives relative to those who reported no.

### Media Exposure to family planning messages

Our findings showed that sources of information on family planning were significantly associated with contraceptive use. Clients who reported a place of worship (aOR=11.97; 95% CI: 1.76-81.62) and community leaders (aOR=6.52; 95% CI: 1.02-41.58) as their source of information on FP showed higher odds of using contraceptives relative to those not exposed to any of these sources. Contrary to expectations, clients that reported receiving family planning messages from a health worker (aOR=0.26; 95% CI: 0.08-0.78) were less likely to use contraceptives when compared to those who did not.

## DISCUSSION

Our study revealed a high prevalence of contraceptive use among the family planning clients, with injectables being the most commonly used method. The high prevalence and pattern of contraceptive use have been supported in previous studies (12,32,33).

Contrary to previous studies,(14,19,34,35) we found no association between quality of FPC and contraceptive use. Although, previous studies have largely associated MII Plus with contraceptive continuation rather than use. However, we found a significant association between other counselling and communication measures with contraceptive use. There is evidence regarding how quality family planning counseling can support women and their partners in adopting contraceptive methods that suit their needs, in addition to assisting them in solving any potential problem that may arise as a result of the use of a particular method(34). Although different contraceptive counseling strategies have been explored in the literature and found to improve contraceptive use and reduce unmet needs. However, there seems to be no clear consensus regarding the best approach to deliver family planning counseling that will be effective in improving contraceptive uptake, reduce unmet need and improve overall client satisfaction(36). This suggest gap in evidence, with further investigation required.

Furthermore, in contrast with most studies (11,29,37) our findings showed exposure to family planning messages was not significantly associated with the use of contraceptives. This may be connected with the low exposure to family planning messages among the women. Although a few studies(24,30) have also reported a non-significant effect of media exposure. There is evidence in literature on how effective mass media can promote accurate information about contraceptives, stimulating attitudinal and behavioural changes, increasing community awareness and mobilization and combating the spread of misinformation and socio cultural and religion myths regarding the use of contraceptives (38–42).

Our study found sources of information about family planning were significantly associated with contraceptive use. Receiving information about family planning from a place of worship and a community leader was significantly associated with the likelihood of using contraceptives. These findings were consistent with previous studies(9,13,23,37) and further reiterate the important roles played by religious institutions and community leaders in promoting the use of modern contraceptives. This is particularly important within the study context because the use of contraceptives is fundamentally shaped by religious and cultural factors.

In line with previous studies(43–45), our findings revealed that individual-level factors, namely literacy level (being able to read or write), currently working, and having autonomy, were significantly associated with the use of contraceptives after adjusting for other factors. Understandably, being able to read or write might allow for a better understanding of counselling information received from the FP provider, especially when communication is a barrier(46). Similarly, active engagement in economic activity relative to being unemployed and being able to jointly decide on contraceptive use with their partner suggests some level of economic and reproductive health empowerment. These no doubt enhances the prospect of using contraceptives among women.

The strength of this study is sampling participants from clients visiting health facilities to access contraceptives, which helped to reduce possible recall bias since questions were asked about their last visit. Regardless, there are a few limitations such as the cross-sectional design, which limited ability to infer a causal relationship, coupled with the limited sample size might have possibly influenced our study outcome. Regardless of these limitations, our findings were in most cases consistent with previous findings, in addition to making important contributions to literature.

## Conclusion

The low quality of family planning counseling and poor exposure to family planning messages suggest the need for developing strategies that will help in strengthening QOC and exposure to multiple sources of information about family planning. This can help to promote contraceptive use. The outcome of this study further echoes the important role played by religious and community leaders in championing and promoting positive social norms toward achieving enhanced modern contraceptive uptake.

## Data Availability

All data produced in the present study are available upon reasonable request to the authors

## Acknowledgement

The authors would like to thank the Bill and Melinda Gates Foundation for funding the study. Also, all the study participants and stakeholders across all the six states where the study was conducted are well appreciated for their support during the research work.

## REFERENCES

1. NBS and UNICEF. Multiple Indicator Cluster Survey 2021, Survey Findings Report. Abuja, Nigeria: National Bureau of Statistics and United Nations Children’s Fund. [Internet]. Abuja; 2022. Available from: https://mics-surveys-prod.s3.amazonaws.com/MICS6/West%20and%20Central%20Africa/Nigeria/2021/Snapshots/Nigeria%20MICS%202021%20Statistical%20Snapshots_English.pdf

2. NPC and ICF. Nigeria Demographic and Health Survey 2018. Abuja, Nigeria, and Rockville, Maryland, USA: NPC and ICF. Abuja, Nigeria, and Rockville, Maryland, USA; 2019.

3. Akpoti OO, Kareem AJ, Kareem AO, Ogunromo AY, Owoeye-Lawal OT, Ahmed LA, et al. Demand for modern contraceptives and use among women of reproductive age in north central Nigeria. Int J Community Med Public Health. 2023 Feb 28;10(3):958–67.

4. Alabi O, Odimegwu CO, Dewet N, Akinyemi JO. Alabi, O., Odimegwu, C. O., De-Wet, N., & Akinyemi, J. O. (2019). Does female autonomy affect contraceptive use among women in northern Nigeria?. African Journal of Reproductive Health. 2019;23(2):92–100.

5. Francis CS, Ahmed AM. Attitude and usage of Contraceptives among Married Couples in Northern Nigeria: A Review. Asian Res J Arts Soc Sci. 2021 Jul 12;25–33.

6. Okafor KC, Omeiza DV, Idoko LO, Inyangobong EA, Unubi OE, Bassi AP. Attitude, Practice, and Factors Affecting Contraceptive Use among Women Attending Postnatal Care in a Tertiary Health Facility in Jos North LGA, Plateau State, Nigeria. Open J Obstet Gynecol. 2022;12(08):814–31.

7. Oladosun M, George TO, Onwumah A, Olawole-Isaac A, Adekoya DO. Modern contraceptive use in Northwestern Region of Nigeria: Rural-Urban segmentation analysis. Afr Popul Stud [Internet]. 2019 Nov [cited 2023 May 15];33(2). Available from: https://aps.journals.ac.za/pub/article/view/1431

8. Sinai I, Anyanti J, Khan M, Daroda R, Oguntunde O. Demand for Women’s Health Services in Northern Nigeria: A Review of the Literature. Afr J Reprod Health. 2017 Jun 30;21(2):96–108.

9. Atama CS, Okoye UO, Odo AN, Odii A, Okonkwo UT. Belief System: A Barrier to the Use of Modern Contraceptives among the Idoma of Benue State, North Central Nigeria. J Asian Afr Stud. 2020 Jun;55(4):600–16.

10. Farrell M, Masquelier A, Tissot E, Bertrand J. Islam, Polygyny and Modern Contraceptive Use in Francophone sub-Saharan Africa. Afr Popul Stud [Internet]. 2014 Sep [cited 2023 May 15];28(3). Available from: http://aps.journals.ac.za/pub/article/view/631

11. Oluwasanu MM, John-Akinola YO, Desmennu AT, Oladunni O, Adebowale AS. Access to Information on Family Planning and Use of Modern Contraceptives Among Married Igbo Women in Southeast, Nigeria. Int Q Community Health Educ. 2019 Jul;39(4):233–43.

12. Adebowale AA, Deborah OI, Olufunke AM, Damilola AO. Prevalence and Pattern of Modern Contraceptive Choices among Women of Reproductive Age 15-49 Years in a Community Health Facility: An Eight Year Retrospective Study. Journal of Advances in Medicine and Medical Research. 2021;33(5):22–3.

13. Prettner K, Strulik H. It’s a Sin-Contraceptive Use, Religious Beliefs, and Long-run Economic Development. Rev Dev Econ. 2017 Aug;21(3):543–66.

14. Jain A, Aruldas K, Tobey E, Mozumdar A, Acharya R. Adding a Question About Method Switching to the Method Information Index Is a Better Predictor of Contraceptive Continuation. Glob Health Sci Pract. 2019 Jun 24;7(2):289–99.

15. Leon FR, Vernon R, Martin A, Bruce L. The Balanced Counseling Strategy: A toolkit for family planning service providers. Washington, DC: Population Council; 2008.

16. Sapkota S, Rajbhandary R, Lohani S. The Impact of Balanced Counseling on Contraceptive Method Choice and Determinants of Long Acting and Reversible Contraceptive Continuation in Nepal. Matern Child Health J. 2017 Sep;21(9):1713–23.

17. Jain A, Aruldas K, Mozumdar A, Tobey E, Acharya R. Validation of Two Quality of Care Measures: Results from a Longitudinal Study of Reversible Contraceptive Users in India. Stud Fam Plann. 2019 May 23;sifp.12093.

18. Palinggi RS, Moedjiono AI, Suarayasa K, Masni, Seweng A, Amqam H, et al. The effect of balanced counseling strategy family planning against attitude, subjective norm, and intentions on the use of modern contraception behavior in the Singgani Public Health Center work area of Palu city. Gac Sanit. 2021;35:S140–4.

19. Chakraborty A, Mohan D, Scott K, Sahore A, Shah N, Kumar N, et al. Does exposure to health information through mobile phones increase immunisation knowledge, completeness and timeliness in rural India? BMJ Glob Health. 2021 Jul;6(Suppl 5):e005489.

20. Chang KT, Mukanu M, Bellows B, Hameed W, Kalamar AM, Grépin KA, et al. Evaluating Quality of Contraceptive Counseling: An Analysis of the Method Information Index. Stud Fam Plann. 2019 Mar;50(1):25–42.

21. ICF International. “Demographic and Health Survey: Interviewer’s Manual.” Rockville, MD: ICF International. [Internet]. 2015. Available from: https://dhsprogram.com/pubs/pdf/DHSM1/DHS7-Interviewer’s-Manual-EN-12Jun2017-DHSM1.pdf.

22. Agarwal S, Sripad P, Johnson C, Kirk K, Bellows B, Ana J, et al. A conceptual framework for measuring community health workforce performance within primary health care systems. Hum Resour Health. 2019 Dec;17(1):86.

23. Adedini SA, Babalola S, Ibeawuchi C, Omotoso O, Akiode A, Odeku M. Role of Religious Leaders in Promoting Contraceptive Use in Nigeria: Evidence From the Nigerian Urban Reproductive Health Initiative. Glob Health Sci Pract. 2018 Oct 3;6(3):500–14.

24. Ahmed M, Seid A. Association Between Exposure to Mass Media Family Planning Messages and Utilization of Modern Contraceptive Among Urban and Rural Youth Women in Ethiopia. Int J Womens Health. 2020 Sep;Volume 12:719–29.

25. Alawode OA, Okeke SR, Sah RK, Bolarinwa OA. Prevalence and determinants of intention to use modern contraceptives among grand-multiparous women in sub-Saharan Africa. Arch Public Health. 2022 Dec 3;80(1):246.

26. Ghosh R, Mozumdar A, Chattopadhyay A, Acharya R. Mass media exposure and use of reversible modern contraceptives among married women in India: An analysis of the NFHS 2015–16 data. Navaneetham K, editor. PLOS ONE. 2021 Jul 13;16(7):e0254400.

27. Singh I, Shukla A, Thulaseedharan JV, Singh G. Contraception for married adolescents (15–19 years) in India: insights from the National Family Health Survey-4 (NFHS-4). Reprod Health. 2021 Dec;18(1):253.

28. Speizer IS, Corroon M, Calhoun LM, Gueye A, Guilkey DK. Association of men’s exposure to family planning programming and reported discussion with partner and family planning use: The case of urban Senegal. Moyer CA, editor. PLOS ONE. 2018 Sep 25;13(9):e0204049.

29. Sserwanja Q, Turimumahoro P, Nuwabaine L, Kamara K, Musaba MW. Association between exposure to family planning messages on different mass media channels and the utilization of modern contraceptives among young women in Sierra Leone: insights from the 2019 Sierra Leone Demographic Health Survey. BMC Womens Health. 2022 Sep 16;22(1):376.

30. Agbadi P, Eunice TT, Akosua AF, Owusu S. Complex samples logistic regression analysis of predictors of the current use of modern contraceptive among married or in-union women in Sierra Leone: Insight from the 2013 demographic and health survey. Todd CS, editor. PLOS ONE. 2020 Apr 16;15(4):e0231630.

31. Agadjanian V. Religious Denomination, Religious Involvement, and Modern Contraceptive Use in Southern Mozambique. Stud Fam Plann. 2013 Sep;44(3):259–74.

32. Mohammed-Durosinlorun A, Abubakar A, Adze J, Bature S, Mohammed C, Taingson M, et al. Comparison of Contraceptive Methods Chosen by Breastfeeding, and Non-Breastfeeding, Women at a Family Planning Clinic in Northern Nigeria. Health (N Y). 2016;08(03):191–7.

33. Muhammed Z, Maimuna DG. Contraceptive trend in a tertiary facility in North Western Nigeria: A 10-year review. Nigerian Journal of Basic and Clinical Sciences. 2014;11(2):99–103.

34. Dehlendorf C, Henderson JT, Vittinghoff E, Grumbach K, Levy K, Schmittdiel J, et al. Association of the quality of interpersonal care during family planning counseling with contraceptive use. Am J Obstet Gynecol. 2016 Jul;215(1):78.e1-78.e9.

35. Hossain S, Sripad P, Zieman B, Roy S, Kennedy S, Hossain I, et al. Measuring quality of care at the community level using the contraceptive method information index plus and client reported experience metrics in Bangladesh. J Glob Health. 2021 Mar 10;11:07007.

36. Cavallaro FL, Benova L, Owolabi OO, Ali M. A systematic review of the effectiveness of counselling strategies for modern contraceptive methods: what works and what doesn’t? BMJ Sex Reprod Health. 2020 Oct;46(4):254–69.

37. Okigbo CC, Speizer IS, Corroon M, Gueye A. Exposure to family planning messages and modern contraceptive use among men in urban Kenya, Nigeria, and Senegal: a cross-sectional study. Reprod Health. 2015 Dec;12(1):63.

38. Ajaero CK, Odimegwu C, Ajaero ID, Nwachukwu CA. Access to mass media messages, and use of family planning in Nigeria: a spatio-demographic analysis from the 2013 DHS. BMC Public Health. 2016 Dec;16(1):427.

39. Chola M, Hlongwana KW, Ginindza TG. Mapping Evidence Regarding Decision-Making on Contraceptive Use among Adolescents in Sub-Saharan Africa: A Scoping Review. Int J Environ Res Public Health. 2023 Feb 3;20(3):2744.

40. Chambongo PE, Nguku P, Wasswa P, Semali I. Community vaccine perceptions and its role on vaccination uptake among children aged 12-23 months in the Ileje District, Tanzania: a cross section study. Pan Afr Med J [Internet]. 2016 [cited 2022 Nov 2];23. Available from: http://www.panafrican-med-journal.com/content/article/23/162/full/

41. Casey SE, Gallagher MC, Kakesa J, Kalyanpur A, Muselemu JB, Rafanoharana RV, et al. Contraceptive use among adolescent and young women in North and South Kivu, Democratic Republic of the Congo: A cross-sectional population-based survey. Wickramage K, editor. PLOS Med. 2020 Mar 31;17(3):e1003086.

42. Nsanya MK, Atchison CJ, Bottomley C, Doyle AM, Kapiga SH. Modern contraceptive use among sexually active women aged 15–19 years in North-Western Tanzania: results from the Adolescent 360 (A360) baseline survey. BMJ Open. 2019 Aug;9(8):e030485.

43. Aduloju-Ajijola NM, Idogho O, Yusuf F, Muhammed F, Anyanti J. Examining the Determinants of Contraceptive Use in Northern Nigeria [Internet]. In Review; 2020 Sep [cited 2023 May 18]. Available from: https://www.researchsquare.com/article/rs-71531/v1

44. Ahinkorah BO, Budu E, Aboagye RG, Agbaglo E, Arthur-Holmes F, Adu C, et al. Factors associated with modern contraceptive use among women with no fertility intention in sub-Saharan Africa: evidence from cross-sectional surveys of 29 countries. Contracept Reprod Med. 2021 Dec;6(1):22.

45. Alabi MA, Fasasi M, Ihimekpen G, Taofeek TA, Oyinloye T, Adedibu D. Factors associated with differentials in modern contraceptive use among currently married women in Nigeria: Muslim North Versus Muslim South. Western Nigeria Journal of Medical Sciences. 2022;5(2):94–102.

46. Assaf S, Wang W, Mallick L. Quality of care in family planning services in Senegal and their outcomes. BMC Health Serv Res. 2017 Dec;17(1):346.

